# Predicting dimensions of depression from smartphone data

**DOI:** 10.1101/2024.01.08.23300679

**Authors:** Vincent L. Holstein, Samir Akre, Ramona Leenings, Yoonho Chung, Tim Hahn, Justin T. Baker

## Abstract

Depressive disorders are highly prevalent but demand nuanced personalized treatment that traditional approaches in psychiatry cannot address. This gap has prompted a surge of interest in leveraging digital technology, particularly smartphones, for remote monitoring to enhance outpatient care. This study utilizes the BRIGHTEN dataset to construct interpretable prediction models for overall depression severity, measured by PHQ-9, and various depression dimensions using a factor modelling approach.

Our factor model unveils a three-factor solution encompassing mood, somatic, and concentration/psychomotor-related factors. Machine learning models effectively predict both the PHQ-9 scores and individual factors, with feature importance methods analyses underscoring the influence of the PHQ-2 scale and communication-related features. These findings are corroborated by models trained on data subsets.

Through nested multi-level models, we identify between-subject effects for the PHQ-2 and select communication-related features, along with within-subject effects for these features. In summary, this study underscores the robust predictive capacity of ecological momentary assessments and highlights features of potential relevance for future investigations, such as communication-related features. We advocate for future studies to assess the cost-effectiveness and intervention potential of these models.

## Introduction

Depression, a pervasive psychiatric disorder, imposes a substantial global burden, affecting millions with an estimated lifetime prevalence of 20.6% [1]. Projections indicate that by 2030, it will eclipse other contributors, becoming the primary worldwide disease burden [2]. As healthcare systems grapple with the escalating impact of depression, there is a growing demand for advanced monitoring and predictive tools to enhance management and enable timely interventions. Moreover, the need for remote monitoring, potentially decreasing visit frequency, has become imperative in response to the challenges posed by the condition.

Acknowledging the limitations of human judgment, both in terms of costliness and biases, recent focus has shifted towards leveraging computational techniques for quantitative behavior assessment [3, 4].

One avenue gaining significant attention in this pursuit is the utilization of digital devices, often termed digital phenotyping or mobile health. Enabled by the ubiquitous use of smartphones and mobile technologies, it offers novel opportunities for remote monitoring and insightful data collection, providing a nuanced understanding of the daily experiences of individuals contending with depression [5, 6]. A particular focus in this field is the precise prediction of mood states or symptoms among individuals with various psychiatric conditions, offering the potential for timely interventions, targeted care, and increased outpatient management flexibility.

In exploring the heterogeneity of depression, past research has delved into the relationship between different phone-derived features and the construction of prediction models for symptoms and overall depression severity [7]. Significant associations have been identified, such as the link between location data and depression severity, especially in terms of homestay time. Lower phone-derived sociability/communication has been correlated with lower mood, alongside changes in daytime and nighttime activity [8–11]. However, the use of varied measures for these behaviors has resulted in considerable heterogeneity in the signals measured. In addition to passive measures, active methods like Ecological Momentary Assessments (EMAs) have demonstrated a significant relationship with composite depression scores, such as the PHQ-9 [12, 13]). These, too, exhibit heterogeneity in the concepts measured and the questions asked across studies.

Moving beyond univariate association testing, research has ventured into the multivariate prediction of depression status, severity, and specific symptoms. Successful classification of depressed individuals and healthy controls has been achieved with accuracies ranging from 0.60 to above 0.85 and AUC values of approximately 0.8 [14–17]. Predicting individual symptoms has shown varying degrees of success for different symptoms [18, 19]. While attempts to predict overall severity often used composite scores of depression symptoms as targets, reliance on passive measures, without incorporating active measures like EMAs, has been a common trend [8, 20– 22].

Given the heterogeneity in passive data across these studies, a pivotal question arises: How can we identify the most relevant features for future digital phenotyping studies? It is crucial to explore whether different aspects of depression are better predicted by specific sensor signals or active measures. Previous studies examining feature importance found robust predictive power for phone usage, internet usage, EMAs, and, to a lesser degree, GPS-derived features in predicting de-pression severity [19, 22]. Diurnal usage patterns emerged as relevant predictors in the prediction of depression status [23]. However, given the diverse underlying apps used in these studies, significant heterogeneity exists in the sensors and signals collected.

This study addresses these gaps by examining the predictability of different dimensions of depression, utilizing a factor modeling approach to discern the severity of specific domains of depressive symptoms. We also explore feature importance through multiple methods, guiding the selection of features for future studies. Finally, employing a multi-level model for within- and between-subject variance, we assess the utility of different features for modeling approaches. We anticipate that the outcomes of this study will significantly contribute to shaping the landscape of future research in the field of digital phenotyping for depression.

## Methods

### Study Data

For our analysis, we used the BRIGHTEN (Bridging Research Innovations for Greater Health in Technology, Emotion and Neuroscience) study data, a publicly available dataset consisting of two studies: BRIGHTEN-V1 and BRIGHTEN-V2 [24]. Both studies were randomized controlled trials evaluating the effectiveness of digital health apps in improving mood. Eligibility criteria included speaking English or Spanish, being above 18 years of age, owning an iPhone or Android and an Apple iPad 2.0 or newer. A score of >5 on the PHQ-9 or >2 on PHQ-9 item 10 was required for inclusion.

Participants received a baseline screening where the PHQ-9, GAD-7, AUDIT-C and IMPACT Mania and Depression screening were administered. Furthermore, demographic information including age, gender, income, race, education and device type were elicited. Afterwards, participants received weekly PHQ-9 surveys for 4 weeks and then every two weeks along with other scales. PHQ-2 was administered daily and passive data was constantly collected for the study duration of 12 weeks. For further information, we refer to Pratap et al [24].

For our analyses, we used the baseline assessment data with completed questionnaires and demographic information, while for all other modelling, we used study data from each participant with more than 4 weeks of data. The data was averaged over one week to reduce dimensionality and provide data relevant to the corresponding PHQ-9 administration. Detailed demographic information can be found in the supplementary materials.

### Data availability

The data is publicly available and can be accessed via the study portal (www.synapse.org/brighten). Users need to request access, as described under the “Accessing the Brighten Study data” page.

### Factor Analysis

For discovering the factor structure of the PHQ-9 scale the baseline PHQ-9 assessment was used. This provided a set of PHQ-9 scales across individuals that were not used in subsequent analyses. First, we examined the skewness of the various items. It was decided to not remove any items.

To maximize the between-group similarity in the exploratory and confirmatory factor analyses, we employed the anticlustering function from the anticlust package for a similar data split [25].

To conduct parallel analyses on the correlation matrices, we utilized the fa.parallel function within the psych package to identify a potential range of factors to retain [26]. All subsequent analyses were conducted in this range.

Following parallel analysis, we employed the lavaan package to fit all the models, subsequently comparing them based on various model fit statistics [27]. Our metrics for comparison encompassed the Tucker-Lewis index (TLI), comparative fit index (CFI), root mean squared error of approximation (RMSEA), information criteria as well as a comprehensive evaluation of the conceptual solutions.

According to the literature, a robust model fit is suggested by CFI values greater than 0.95 and RMSEA values below 0.05 [28, 29]. Furthermore, an acceptable fit is indicated by a CFI higher than 0.90 and an RMSEA ranging from 0.05 to 0.08.

### Predictive Modelling

All predictive models were implemented in the PHOTONAI Python package [30]. Standard machine learning estimators available in scikit-learn (Linear Regression, Automatic Relevance Determination (ARD) Regression, Support Vector Regressor, Random Forest Regressor, Gradient Boosting Regressor) are already integrated into the PHOTONAI package [31]. The Mixed Effects Random Forest (MERF) was integrated using custom data wrappers in PHOTONAI and is based on the MERF Python package (https://github.com/manifoldai/merf) [32].

To encode missingness additional indicator columns were generated encoding the missingness of the averaged values as either 0 (not missing) or 1 (missing). Missing values in the regular columns were then set to zero. This was done to allow models to use information about missing data information in their prediction.

Each pipeline, consisting of a Standard Scaler (removing the mean and scaling to unit variance) and an estimator was trained in a 10x10 nested cross-validation. Models were selected on mean absolute error using grid search as an optimization strategy. Additionally mean squared error, Spearman correlation and Pearson correlation were calculated, to provide further insight into model performance.

Each pipeline was evaluated in a user-split and random-split scenario, where the stratified folds in the K-Fold split were either based on the user or based on a random shuffling of data points where data points from most subjects were present in the different folds. As baseline estimation is a strong driver of effects we performed both split scenarios to evaluate generalizability with data points present and generalizability to new users.

Each pipeline was compared against a dummy regressor, a model which predicts the mean of the training set in each test set. Furthermore we also compared models to a baseline dummy that predicted the average for each individual present in the training data set and the average across the group for individuals not present in the data set. Average model performance was compared to the dummy models. We also performed permutation tests for each model, to assess whether these models predicted the target variable significantly above chance.

### Feature Importance

To investigate feature importance and how it relates to the model’s decision-making, the best-performing model for each target was selected and Shapley additive Explanation (SHAP) scores were generated for each feature. The kernel explainer from the shap package was used for calculating SHAP scores [33]. These scores help explain individual model predictions giving researchers the opportunity to understand how different features contribute to model predictions.

### Subset Models

Further investigating the importance of subgroups of data we trained predictive models on defined subsets of data, containing data measuring one type of concept. For V1 we subset data to EMA data, communication data and mobility data. For V2 subset data to EMA data, communication data, activity data and mobility data. Each data subset was combined with demographic data and prediction models were trained on these subsets in a user and random split using either an ARD regression or a MERF. A table of the different subsets can be found in the appendix

### Inferential Modelling

The study utilized inferential models to analyze the statistical relationships between participants’ symptoms and various digital measures. Controlling for participant gender, the models incorporated multilevel regression techniques, considering both between-person and within-person components. The varying effects were estimated, allowing for different intercepts and slopes among participants. The results were interpreted based on population-level effects, with standardized slopes calculated for interpretability and comparability. To accommodate skewed outcome variable distributions, the study employed the skew-normal distribution, parameterized to allow for non-zero skewness.

We established prior distributions for our model parameters, aiming to eliminate unreasonable values while allowing for reasonable ones [34]. The six main types of parameters in our skew-normal multilevel regression models included slopes, intercepts, varying effects’ standard deviations, correlations between varying effects, residuals’ standard deviation, and the skewness parameter. To ensure stable estimates and prevent overfitting, we used more conservative normal priors centred on zero for the slope parameters. For the intercept parameters, we employed less conservative Student’s t priors. Nonnegative Student’s t priors centred on zero were utilized for the varying effects’ standard deviations, while the correlations between varying effects followed the approach of Lewandowski et al. to assign equal prior probabilities to all valid correlation matrices [35]. The standard deviation of the residuals adhered to Student’s t priors centred on zero. Additionally, normal priors centred on zero were applied to the skewness parameter.

To interpret our inferential models, we determined each slope’s magnitude using the posterior median and its precision using the 89% highest density interval (HDI). The posterior median minimizes the expected absolute error, while the 89% HDI, chosen for its stability, represents the narrowest continuous interval encompassing 89% of the posterior density [36].

All inferential analyses were conducted using the brms R package utilizing Markov chain Monte Carlo sampling via the No-U-Turn Sampler algorithm [34, 37, 38].

## Code Availability

The code used for this analysis is publicly available on GitHub under https://github.com/VHolstein/brighten_dimensions.

## Results

### Factor Analysis

Our parallel analysis showed observed eigenvalues exceeding the simulated eigenvalues for 2 to 4 factors. In accordance with these results, we developed models for 2 to 4-factor solutions, where only the 2 and 3-factor solutions converged. Comparing model performance both 2 and 3-factor solutions showed a good model fit during EFA (2-factor model: CFI 0.953, RMSEA 0.082, BIC 22228.870; 3-factor model: CFI 0.992, RMSEA 0.041, BIC 22168.222). We estimated a CFA with thresholding of factor loadings (>0.4 and >0.5) again showed a strong performance for the 3-factor model (3-factor 0.5 threshold model: CFI 0.995, RMSEA 0.060, BIC 10261.572).

Due to the strong model performance, We examined the factor loadings for the 3-factor solution, with the first factor reflecting mood-related contents, the second factor reflecting somatic symptoms and the third factor reflecting concentration/psychomotor symptoms. Due to strong model performance and well-aligned factor loadings, we chose the 3-factor model to model factor scores. We calculated factor scores both in the baseline sample used for modelling as well as the study sample used for predictive and inferential modelling.

### Predictive Modelling

To predict the PHQ9 sum score from all available measures, we trained 6 different estimator pipelines on both the V1 (3007 datapoints, 37 features, 541 subjects) and V2 dataset (1159 datapoints, 111 features, 276 subjects) respectively. For more information on missing datapoints per subject, please see the supplementary materials. These pipelines were run under random and user-split conditions described in the methods section.

PHQ9 was best predicted by an ARD Regression model in the user-split scenario both in V1 (Mean absolute error (MAE): 3.041; Pearson correlation: 0.698) and V2 (MAE: 3.625; Pearson correlation: 0.648). In the random split scenario the mixed effects random forest was the best-performing model both in V1 (MAE: 2.216; Pearson correlation: 0.840) and V2 (MAE: 2.386; Pearson correlation: 0.838). The Baseline Dummy performed worse than the MERF in both random- and user-split in V1 (Random-Split MAE: 2.508 User-Split MAE: 4.341) and V2 (Random-Split MAE: 2.409; User-Split MAE: 4.971) These results can be observed in figure 2.

**Fig. 1.**
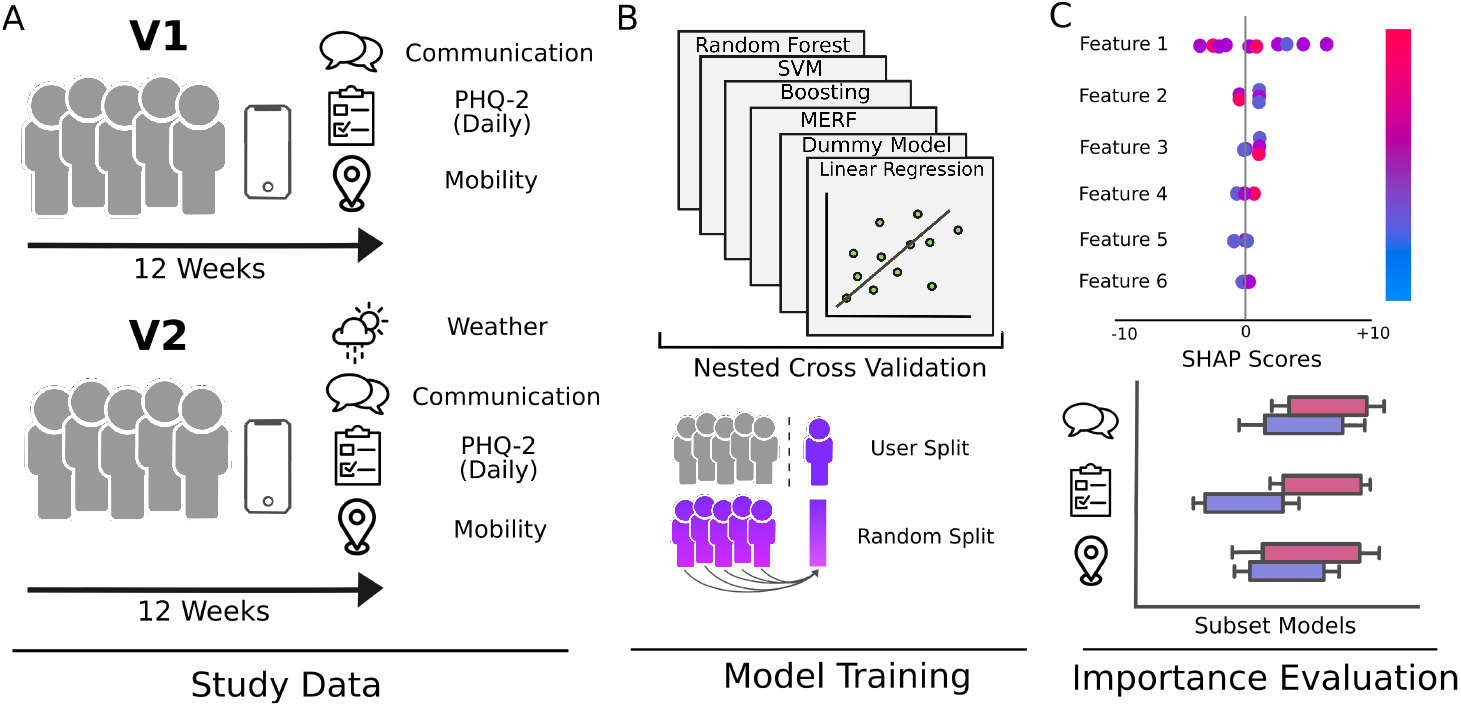
Analysis Overview. A) For all analyses the publicly available BRIGHTEN dataset was used which consists of two studies V1 and V2. Each study ran for 12 weeks, with daily collection of PHQ-2, continuous collection of passive measures and weekly administration of PHQ-9 for 4 weeks and bi-weekly afterwards. B) This dataset was used to build prediction models from multiple estimators which were evaluated in user-split and random-split scenarios. C) The best-performing pipelines were selected for further evaluation with SHAP scores and subset models.

**Fig. 2.**
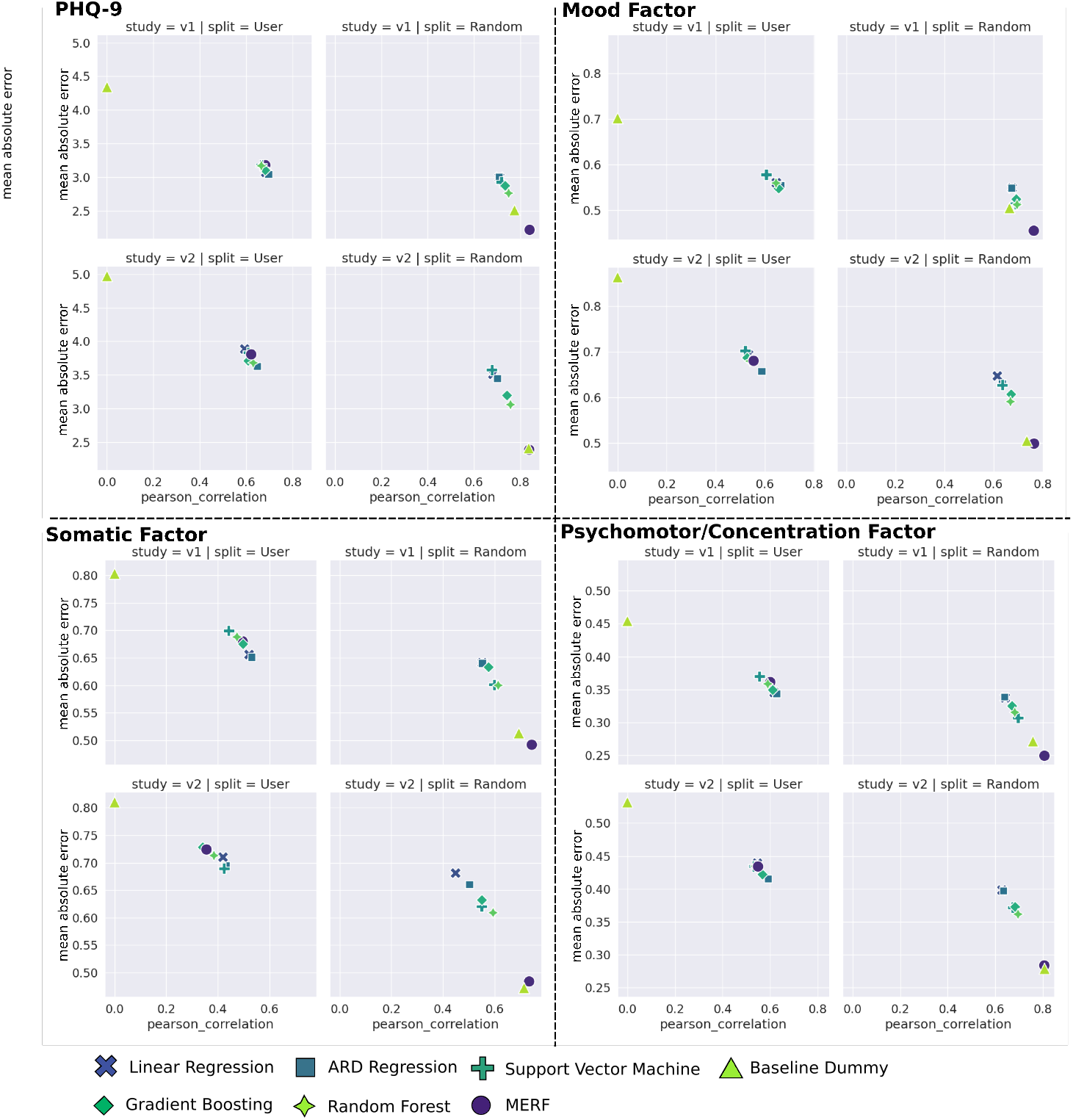
Predicting PHQ-9 Total and Factor Scores. Each quadrant shows the performance of pipelines predicting a specific score. Each plot evaluates the performance of different models in terms of mean absolute error and Pearson correlation for different estimators. The best-performing estimator can be found in the lower right corner of each plot.

**Fig. 3.**
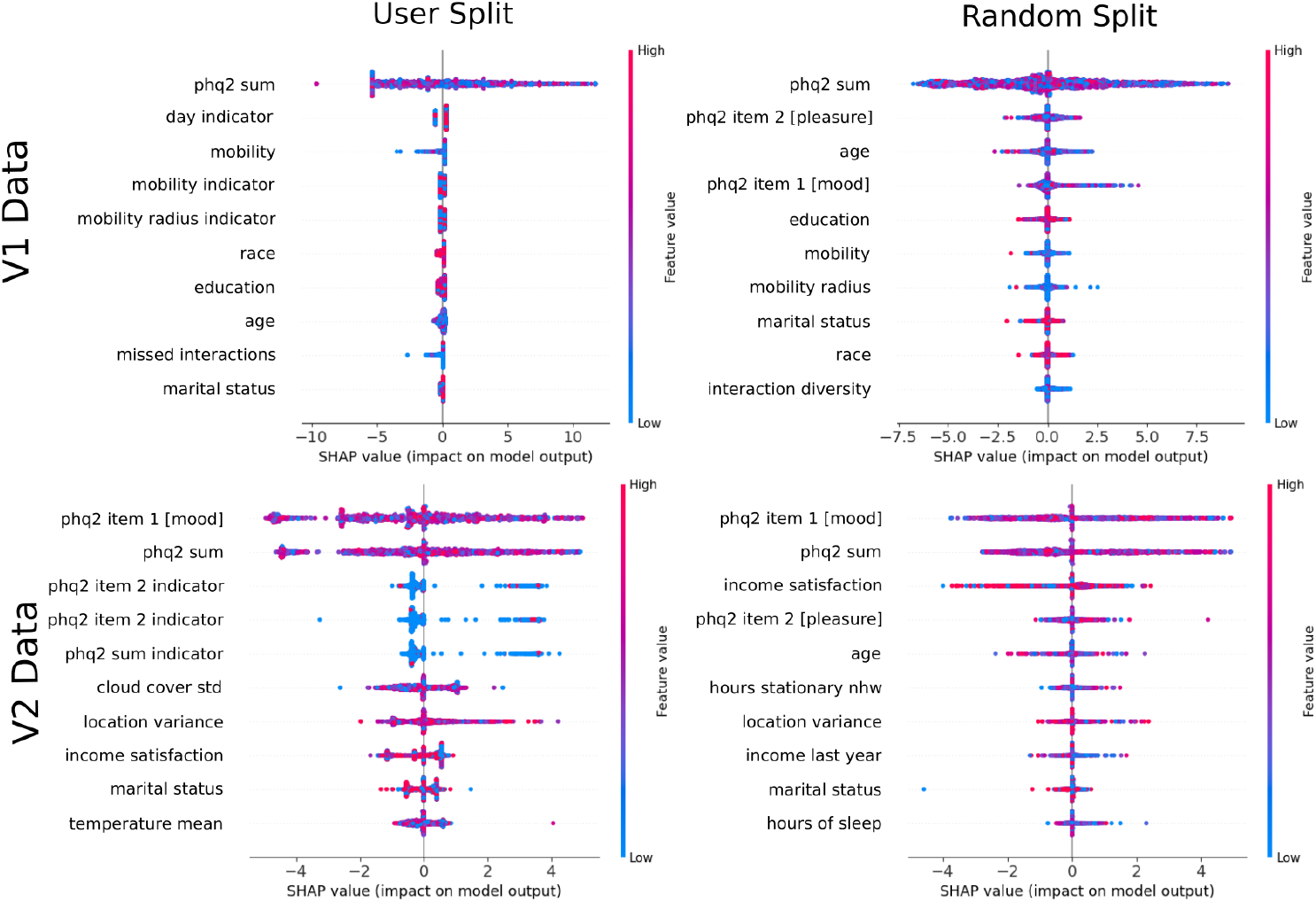
SHAP feature for PHQ9 Prediction. SHAP plots show the top ten features for PHQ9 prediction in V1 and V2 data for user- and random-split. Each dot represents a prediction with color indicating the feature value and the SHAP value indicating the influence of the feature. The SHAP value magnitude indicates the importance for model predictions.

When predicting mood factor scores the Gradient Boosting model performed best in the user-split scenario for V1 (Mean absolute error: 0.548; Pearson correlation: 0.657) while the ARD regression performed best for V2 (Mean absolute error: 0.656; Pearson correlation: 0.589). For the random split, the MERF again was the best-performing model in V1 (Mean absolute error: 0.455; Pearson correlation: 0.764) and V2 (Mean absolute error: 0.498; Pearson correlation: 0.765). The baseline dummy performed worse than the MERF in V1 (Random-Split MAE: 0.505; User-Split MAE: 0.702) and V2 (Random-Split MAE: 0.472; User-Split MAE: 0.863).

When predicting somatic factor scores the ARD regression performed best in the user-split scenario for V1 (MAE: 0.651; Pearson correlation: 0.532) while the Support Vector regression (SVR) performed best for V2 (MAE: 0.689; Pearson correlation: 0.425). The MERF performed best in the random split scenario for V1 (MAE: 0.492; Pearson correlation: 0.744) and V2 (MAE: 0.484; Pearson correlation: 0.734). The baseline dummy outperformed the MERF in V2 in a random-split setting MAE: 0.472), and was beaten by models in all other scenarios.

Concentration/Psychomotor factor scores were best predicted by the ARD regression in the user-split scenario for V1 (MAE: 0.343; Pearson correlation: 0.627) and V2 (MAE: 0.415; Pearson correlation: 0.592). The MERF predicted best in the random split scenario (V1: MAE: 0.249, Pearson correlation: 0.806; V2: MAE: 0.284, Pearson correlation: 0.806). The baseline dummy outperformed the MERF in V2 in a random-split scenario (MAE: 0.278) and was again beaten in by the models in all other scenarios.

### Feature Importances

SHAP scores were calculated based on ARD Regression in a user split, and the MERF in the random split scenario. The ten most important features from all folds were investigated for predicting the PHQ-9 total score and each of the factors in V1 and V2.

Using SHAP scores to estimate feature importance in V1 revealed PHQ-2 sum score, mobility and missed interactions along with demographic variables as the most important features related to PHQ-9 prediction. Similar patterns were observed for the mood factor, the somatic factor and the psychomotor/concentration factor in the user split scenario. In a random-split scenario, the most important variables were the PHQ-2 sum score, PHQ-2 items 1 and 2, mobility, mobility radius and interaction diversity as important features. Similar patterns were evident for the three factor scores.

Using SHAP scores to investigate feature importance in V2 revealed PHQ-2 sum score, PHQ-2 item 1, cloud cover standard deviation, location variance, temperature mean and demographic variables as the most important features related to PHQ-9 prediction. For the mood factor, somatic factor and psychomotor/concentration factor we observed similar patterns regarding PHQ-2 with slight differences in the most important passive features. In a random-split scenario the PHQ-2 scores emerged as the most important predictors along with hours stationary, location variance, hours of sleep and demographic variables. As before PHQ-2 effects were consistently present across factor scores with considerable variability regarding the most important passive features.

### Subset Models

After examining feature importances we also evaluated model performance for models trained on data subsets based on different data types.

In V1 models based on EMA outperformed Dummy models in both user- () and random-split () scenarios, while MERF models based on communication or mobility outperformed Dummy models in the random split.

In V2 models based on EMA outperformed Dummy models in both user and random split, while MERF models based on communication, activity or weather data outperformed Dummy models in the random split. ARD regression models performed slightly better or comparably to the Dummy estimator.

### Inferential Modelling

For the PHQ9 total score and each of the factor scores, we first tested for associations with the different digital measures in both V1 and V2. The results for PHQ9 total are depicted in Fig. 4. In V1 between-person slopes were significant for all EMA responses, interaction diversity, missed interactions and unreturned calls: increased use of PHQ2 sum score associated with higher PHQ9 (B=0.775, pd=100%), as did higher PHQ Item 1 (B=0.772, pd=100.0%) and Item 2 response (B=0.0.767, pd=100.0%). Increased interaction diversity (B=0.154, pd=98.3%), increased missed interactions (B=0.148, pd=98.5%) and increased unreturned calls (B=0.162, pd=99.6%) all associated with a higher PHQ-9. For EMA responses similar patterns were observed across all factors, while for interaction diversity and missed interactions were significantly associated with higher psychomotor/concentration factor scores only. Unreturned calls were significantly associated with the mood (B=0.129, pd=97.7%) and psychomotor/concentration factor (B=0.151, pd=99.4%).

**Fig. 4.**
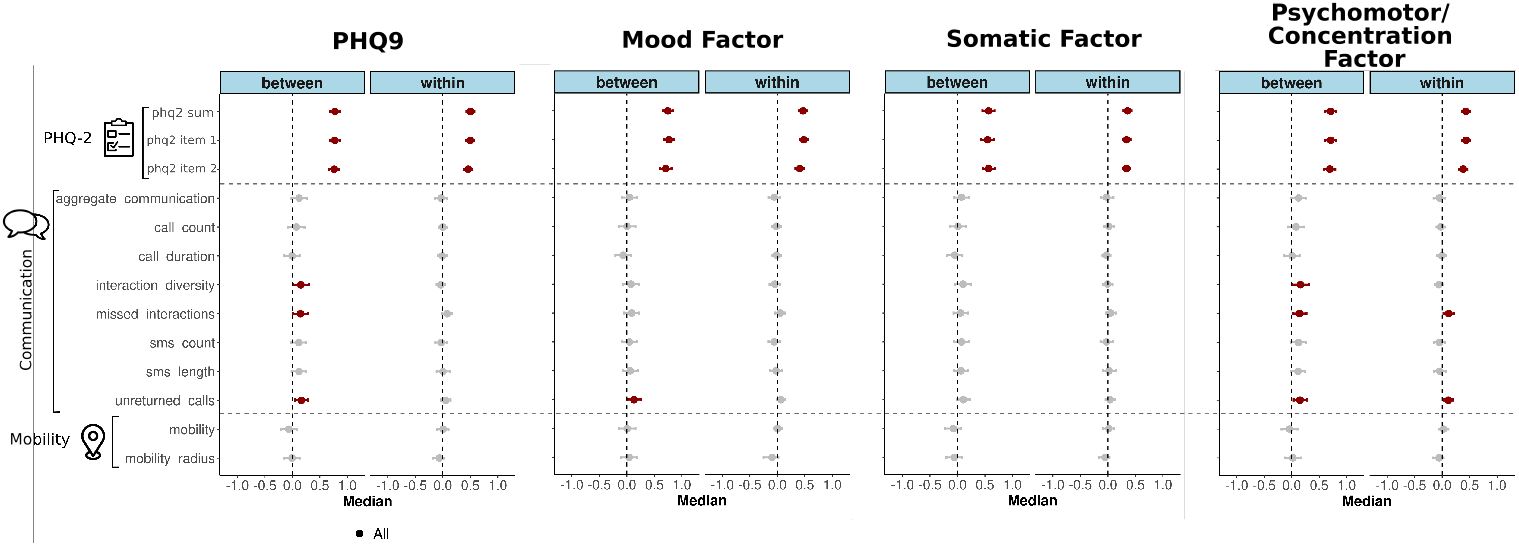
Between- and within-subject importance in V1. Forest plot showing the standardized slope estimates in the inferential models for the PHQ-9 and each of the factor scores in V1. Points represent posterior medians and intervals are posterior 95% highest density intervals. Significant associations are coded in red, and non-significant associations are in grey.

**Fig. 5.**
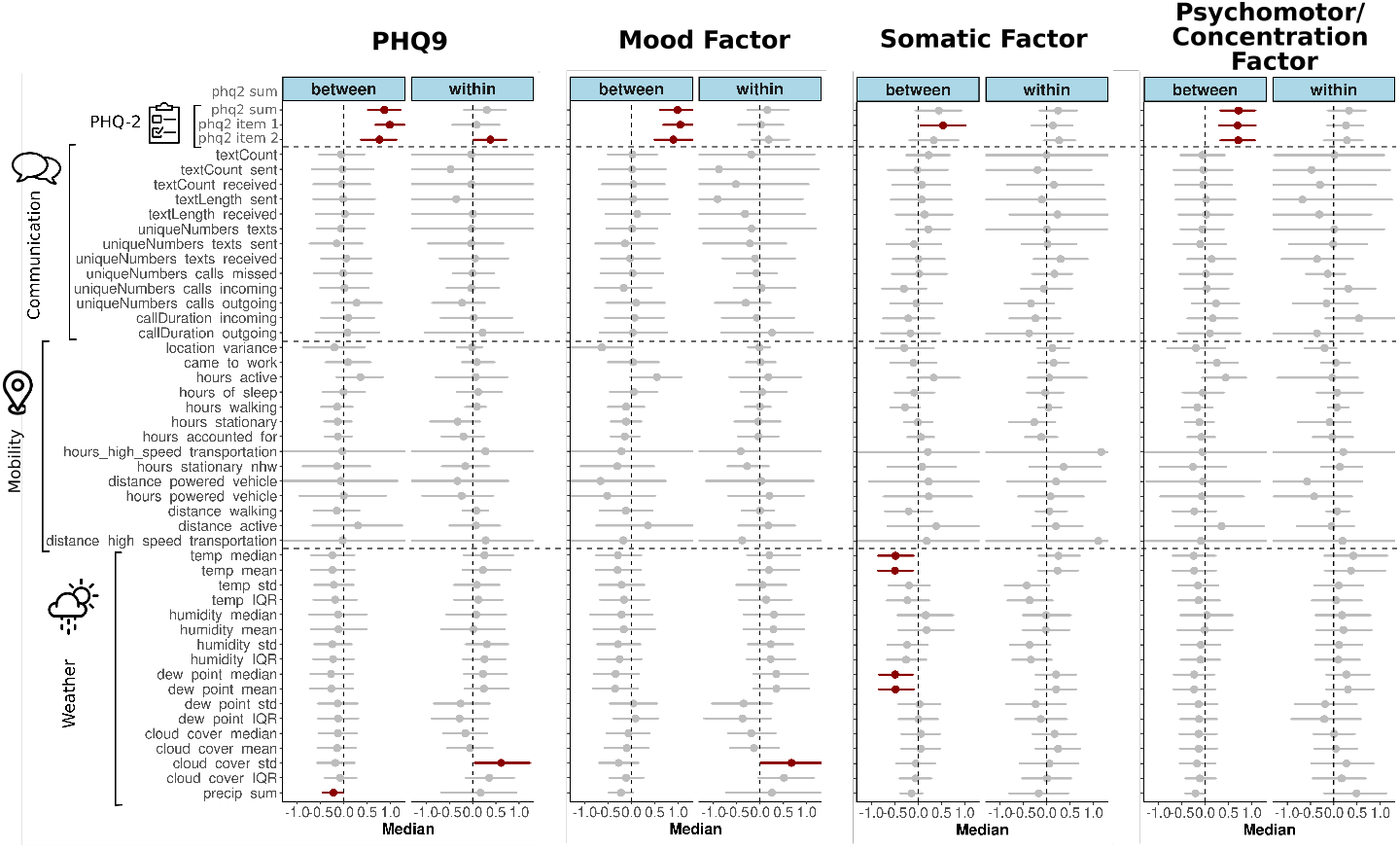
Between- and within-subject importance in V2. Forest plot showing the standardized slope estimates in the inferential models for the PHQ-9 and each of the factor scores in V2. Points represent posterior medians and intervals are posterior 95% highest density intervals. Significant associations are coded in red, and non-significant associations are in grey.

**Fig. 6.**
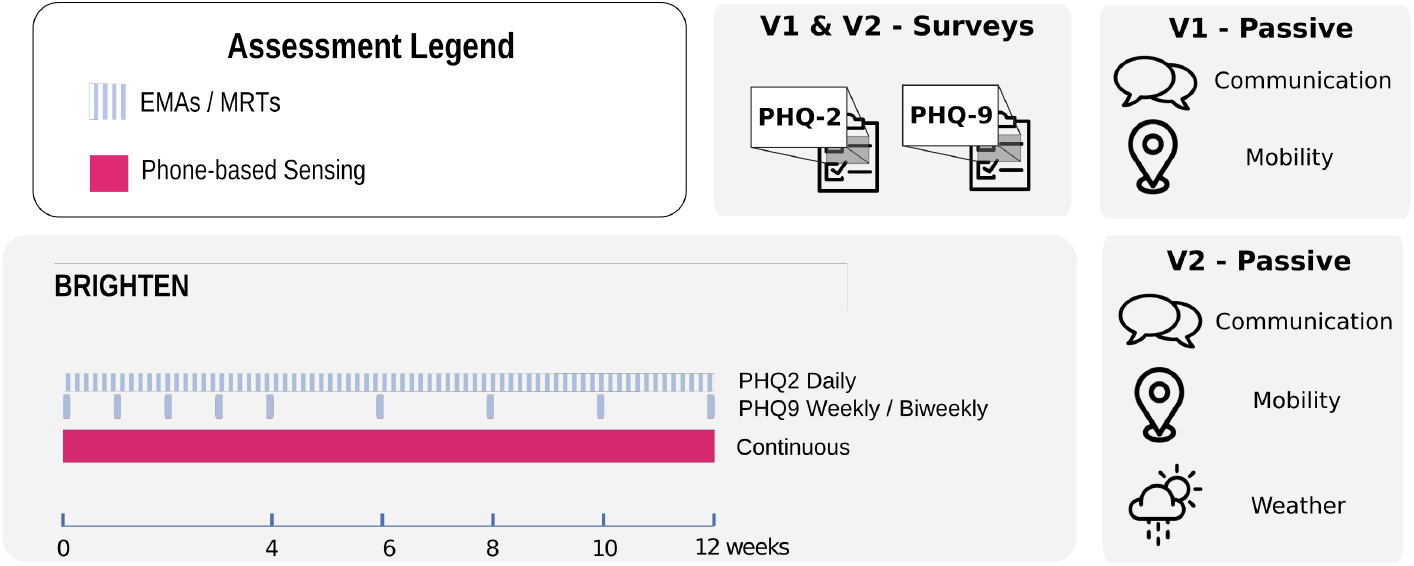
BRIGHTEN Study protocol.

**Fig. 7.**
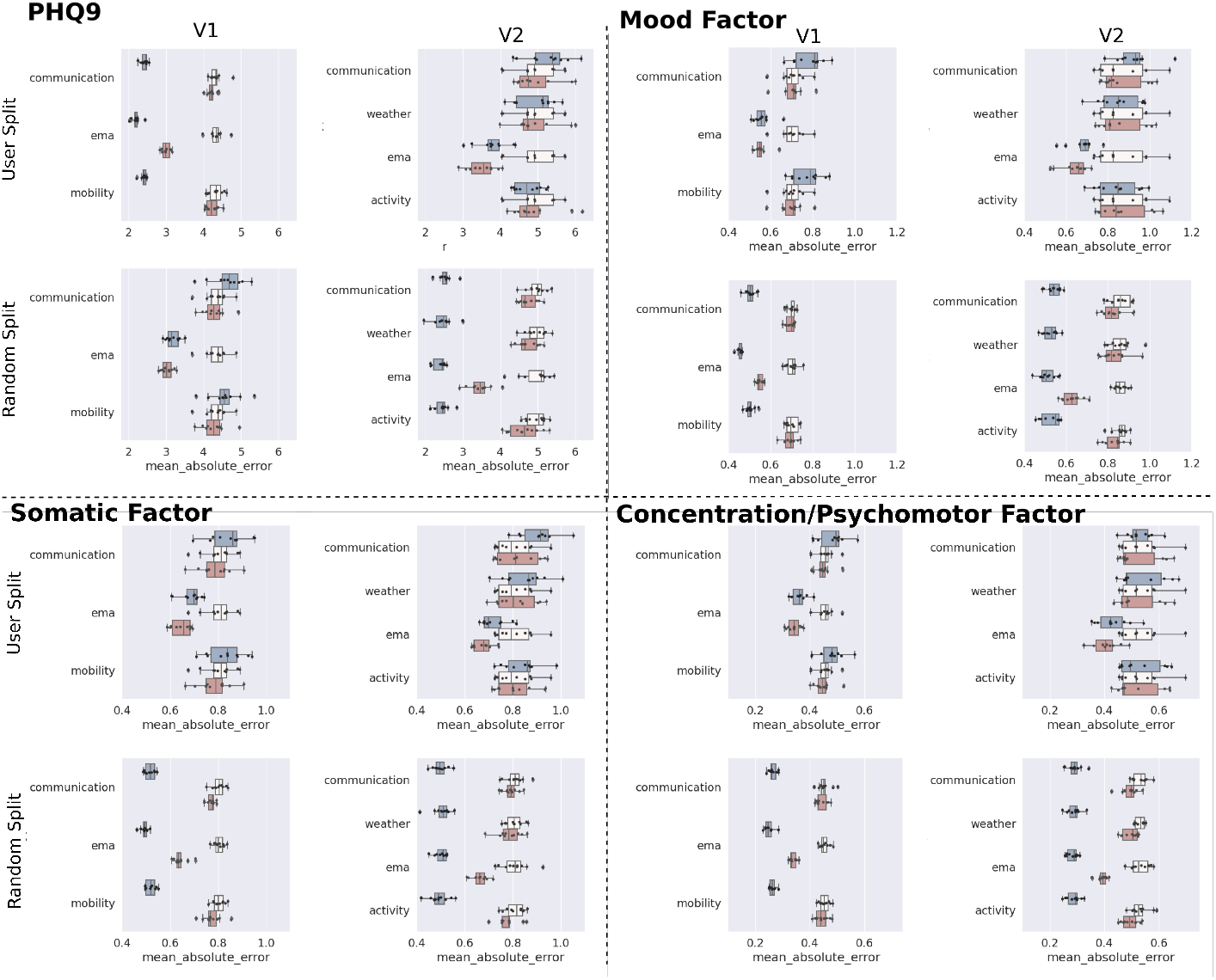
Subset Model Performance for PHQ-9 Total Score

Within-person slopes were significant for all EMA responses, missed interactions and unreturned calls: PHQ2 sum score (B=0.503, pd=100.0%), PHQ2 item 1 (0.500, pd=100.0%) and PHQ2 item 2 (B=0.462, pd=100.0%) were associated with higher PHQ-9 scores and each of the three factors. Increased unreturned calls (B=0.114, pd=99.6%) and missed interactions (B=0.120, pd=99.3%) were associated with increased psychomotor/concentration factor scores, but not PHQ-9 or the other factors.

In V2 between-person slopes were significant for PHQ-2 sum score, PHQ-2 item 1, PHQ2 item 2 as well as temperature mean/median and dew point mean/median. Higher PHQ-2 sum score (B=0.868, pd=99.9%) is associated with higher PHQ-9, as are PHQ-2 item 1 (B=0.983, pd=100.0%) and item 2 (B=0.762, pd=99.9%). Similar patterns were observed for factors 1 and 2. Lower temperature mean (B=-0.497, pd=99.1%), lower temperature median (B=-0.485, pd=99.1%), lower dew point mean (B=-0.485, pd=99.1%) and lower dew point median (B=-0.494, pd=99.4%) were associated with higher factor 2. Within-person slopes in V2 were not significant in any of the predictors.

## Discussion

In summarizing the results of our analysis, we examined various sub-factors of depression and their predictability using phone data. We conducted an extensive examination, encompassing the relevance of different features through feature importance analysis and subset models. Additionally, we explored the impact of different predictors using multi-level models to discern between- and within-subject effects.

Examining the dimensions of depression severity, our initial factor analysis suggested a 3-factor structure for the PHQ-9 scale, aligning with previous findings of a 3-factor structure as part of a hierarchical factor solution [39]. Our three factors reflected mood-related content, somatic symptoms and concentration/psychomotor symptoms, closely aligning with the results by Guerra et al (somatic, cognitive/affective, concentration/motor). Ohter papers often reported a two-factor structure, however these often did not evaluate a three-factor solution [40–42]. Our findings underscore the nuanced nature of depression, highlighting the importance of considering multiple dimensions in remote assessments.

Utilizing diverse machine learning estimators for the prediction of depression severity measured by PHQ-9, we achieved robust and consistent predictions across user- and random-split scenarios. Each model demonstrated performance significantly above chance, as confirmed by permutation tests. In the random-split scenario, the mixed random forest emerged as the superior model in V1 and for overall PHQ-9 and mood factor prediction in V2, aligning with findings from Lewis et al. [21]. However, in V2, the MERF was surpassed by the baseline dummy for somatic and psychomotor/concentration scores, consistent with the weaker inferential significances for multiple features in V2. In the user-split scenario, ARD regression predominantly outperformed other methods, suggesting that baseline estimation via random effects strongly influenced MERF performance. Our model’s observed performance aligns with previous studies predicting PHQ-9 or other depression severity measures on a continuous scale [8, 20, 21]. Notably, our use of averaged data to prevent overfitting distinguishes our approach from some other machine learning studies that employed a more finegrained temporal resolution [7, 8]. An important avenue for future exploration involves comparing the utility of different distributional summaries, such as median, mean, and standard deviation as features in prediction models.

Evaluating the feature importance of our models, the pre-eminent role was attributed to the PHQ-2, marked by consistently high importance across different models as indicated by SHAP scores. This importance was indicated for both the PHQ-2 summary score and its individual items, with variation between the different models. Additionally, communication-related features, encompassing missed interactions, along with location-related features, and in V2, weather-related features, demonstrated a moderate level of importance in both split conditions. Notably, V2 exhibited more considerable variability than V1, aligning with observations from previous univariate studies that hint at a correlation between depression severity and communication-derived and location-derived features [9, 43]. Moreover, our findings reinforce the robust predictive power of Ecological Momentary Assessments (EMAs) in discerning depression trends [22].

The consistency of these trends was further substantiated by various subset models. Models grounded in the PHQ-2 consistently outperformed Dummy models across both user- and random-split conditions. In contrast, models relying on passive data exhibited similarities to Dummy models in both splits, albeit with a slight improvement on average. Notably, the MERF demonstrated superior performance in the user-split scenario, potentially attributable to its described baseline estimation through the random intercept.

To gain a deeper understanding of the relationships between different predictors and our targets, we employed nested multi-level models to examine both between- and within-subject effects. Our inferential approach not only affirmed prior observations regarding the robust predictive power of the PHQ-2 but also revealed subtle between-subject effects for communication-related features in V1 and certain weather-related items in V2. This corroborates findings from earlier studies, suggesting that communication-related features may play a significant role in influencing both between- and within-person effects [7, 9]. The observed association with weather patterns, differing from previous results, underscores the potential impact of geographical het-erogeneity, as earlier studies were conducted in more homogenous regions [44]. It’s essential to note a limitation in this study’s communication-derived features, characterized by limited availability and a high degree of missingness. Future studies with denser availability of these features may enhance predictive capabilities.

Collectively, these findings underscore a robust influence of the PHQ-2, akin to a form of Ecological Momentary Assessment (EMA), while also revealing more modest yet persistent effects for communication-related items. This aligns coherently with earlier research indicating comparatively subdued impacts of passive data [21, 22].

These outcomes provide additional evidence for the formidable predictive capabilities of EMAs, prompting a crucial inquiry: What precisely is the role of passive data in remote monitoring, and what data streams/features do we need to collect?

Our findings suggest that valuable insights can be derived from measures of communication, and employing more advanced metrics could yield enhanced features in the realm of remote monitoring. For instance, exploring the sentiment of sent messages could be highly promising as linguistic features might offer an even more precise understanding of the current mood state, as evidenced in studies exploring sentiment analysis in other contexts [45, 46]. Additionally, our results present some evidence supporting the utility of weather-related features, which can be conveniently gathered in studies leveraging GPS signals. Here further investigation is necessary to determine the utility of these signals.

A pivotal challenge for forthcoming studies leveraging phone data lies in the imperative to ensure comprehensive and precise data sampling. In this investigation, we employed indicator variables to capture missing data and maintain sample size; however, more densely sampled data holds promise for improving prediction accuracy. The choice of sampled data in subsequent studies will hinge significantly on the target disease and the specific outcome measure. Our inferential analyses unveiled the impact of communication variables on the PHQ-9 scale and the psychomotor/concentration factor, though this effect did not extend to other factors in V1. To validate and generalize these findings, further research is warranted, emphasizing the necessity for future studies to carefully consider the specific outcome of interest.

A prospective application of passive data involves determining a threshold, beyond which an Ecological Momentary Assessment (EMA) is triggered to gather supplementary information. This approach leverages passive data to mitigate alert fatigue that may arise from recurrent EMA responses, offering a judicious balance between timely intervention and the associated costs or non-intervention. Identifying optimal cut-offs necessitates a careful consideration of alert fatigue, intervention costs, and the potential implications of non-intervention, thereby facilitating an accurate assessment of depression severity when required.

This limitation is integral to the overall scope of our analysis. While our primary focus centered on predicting either PHQ-9 total scores or dimensions (factors) of depression, we did not explicitly evaluate the advantages of these predictions. To conduct a more comprehensive assessment, a cost-benefit analysis would be imperative, particularly concerning potential interventions, mirroring approaches employed in prior studies on suicidality prediction [47]. This analysis could be complemented with tailored prediction algorithms for specific symptoms necessitating immediate attention, such as suicidality. Although earlier attempts have been made to predict individual symptoms from phone data [18, 19], these analyses predominantly focused on binary outcomes rather than the nuanced severity of individual symptoms, with the added nuance of overlooking cost-benefit considerations.

In conclusion, this analysis provides compelling evidence underscoring the considerable promise of smartphone data in the remote monitoring of depression across diverse dimensions of severity. Our findings notably highlight the robust predictive capabilities of Ecological Momentary Assessments (EMAs), contributing valuable insights into the potential utility of certain passive features. These insights, gleaned from our exploration, offer a foundation for shaping future studies in remote device monitoring for depression. Moreover, our discussion delves into prospective avenues for advancing this line of research, emphasizing key questions that warrant exploration in forthcoming studies.

## Data Availability

All data used in the manuscript are publicly available under: www.synapse.org/brighten

## ACKNOWLEDGEMENTS

Data in this analysis was taken from the publicly available BRIGHTEN dataset (www.synapse.org/brighten).

## Supplementary Material

|  |  |
| --- | --- |
| Female | 1429 |
| Male | 490 |
| Community_College | 412 |
| Elementary_School | 6 |
| Graduate_Degree | 310 |
| High_School | 439 |
| None | 2 |
| University | 750 |
| NoWork | 619 |
| YesWork | 1300 |
| MarriedOrPartner | 631 |
| Separated | 213 |
| Single | 1075 |
| Black | 186 |
| Native | 17 |
| Asian | 130 |
| MultipleRace | 57 |
| White | 1057 |
| Hispanic | 458 |
| PacificIslander | 5 |
| OtherRace | 9 |
| Android | 528 |
| IPhone | 1391 |
| Age_Range | 18-76 |
| Age_min | 18 |
| Age_max | 76 |
| Age_Median | 30 |
| Age_StdDev | 11.090308700715573 |

**Factor Analysis**.EFA Table

CFA Table

Missingness Table

Model Performance

Submodels

**Table 1.**
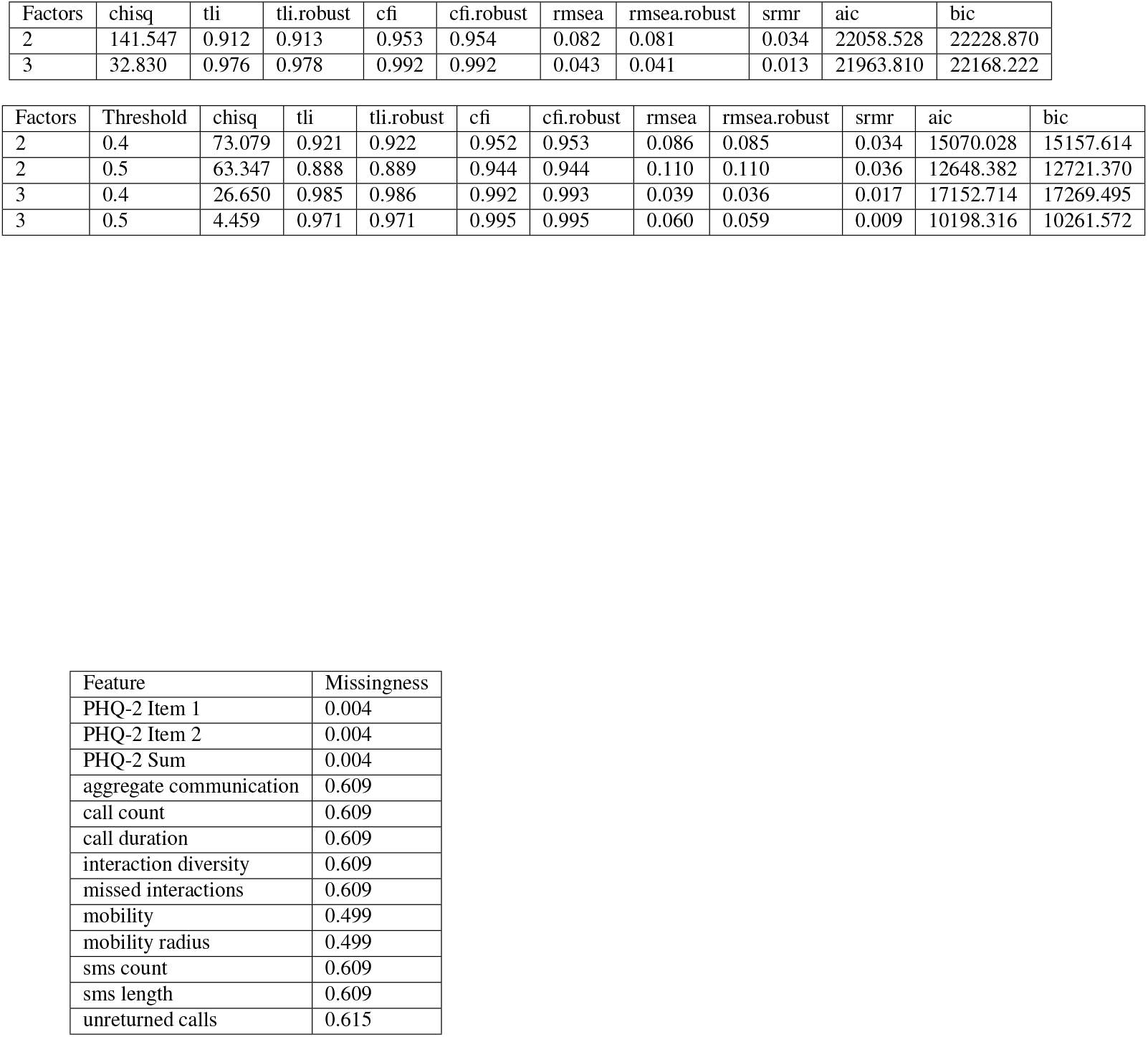
Missingess in V1 as a fraction of total collection points.

**Table 2.** Missingness in V2 as a fraction of total collection points.

|  |  |
| --- | --- |
|  | 0 |
| PHQ-2 Item 2 | 0.113 |
| cloud cover std | 0.356 |
| distance powered vehicle | 0.356 |
| text Length received | 0.933 |
| temp median | 0.356 |
| text Count sent | 0.933 |
| dew point median | 0.356 |
| cloud cover median | 0.356 |
| dew point mean | 0.356 |
| unique Numbers calls missed | 0.933 |
| humidity std | 0.356 |
| precip sum | 0.356 |
| hours powered vehicle | 0.356 |
| cloud cover mean | 0.356 |
| call Count missed | 0.933 |
| dew point IQR | 0.356 |
| textLength sent | 0.933 |
| hours high speed transportation | 0.356 |
| hours accounted for | 0.356 |
| hours stationary nhw | 0.356 |
| unique Numbers calls outgoing | 0.933 |
| hours stationary | 0.356 |
| text Count received | 0.933 |
| location variance | 0.357 |
| call Count outgoing | 0.933 |
| distance walking | 0.356 |
| unique Numbers texts received | 0.933 |
| PHQ-2 Sum | 0.113 |
| hours walking | 0.356 |
| temp mean | 0.356 |
| distance active | 0.356 |
| PHQ-2 Item 1 | 0.113 |
| hours of sleep | 0.356 |
| call Duration outgoing | 0.933 |
| temp std | 0.356 |
| temp IQR | 0.356 |
| call Count incoming | 0.933 |
| humidity median | 0.356 |
| text Count | 0.933 |
| unique Numbers calls incoming | 0.933 |
| unique Numbers texts | 0.933 |
| came to work | 0.345 |
| humidity IQR | 0.356 |
| distance high speed transportation | 0.356 |
| unique Numbers texts sent | 0.933 |
| humidity mean | 0.356 |
| call Duration incoming | 0.933 |
| hours active | 0.356 |
| dew point std | 0.356 |
| cloud cover IQR | 0.356 |

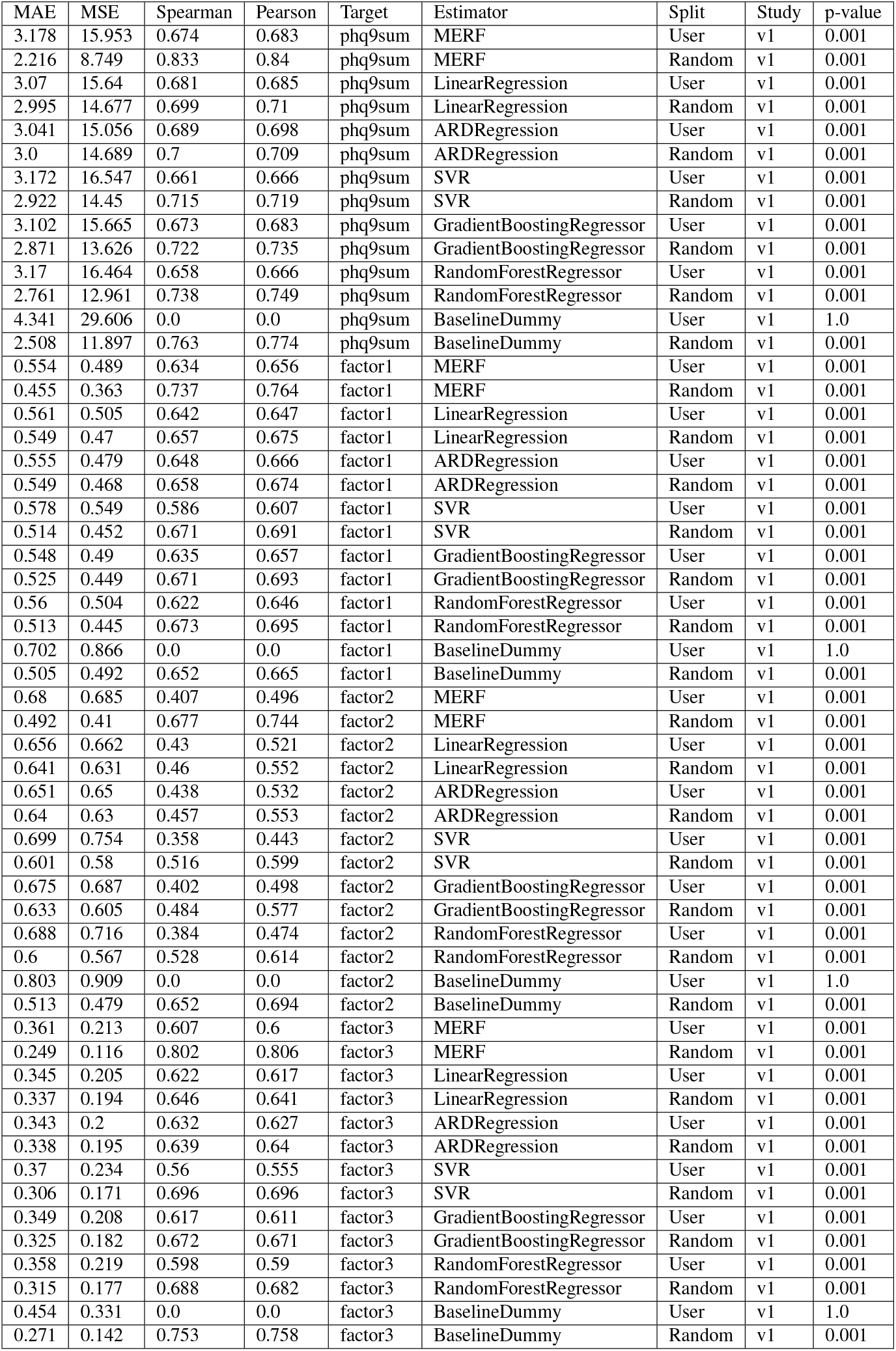
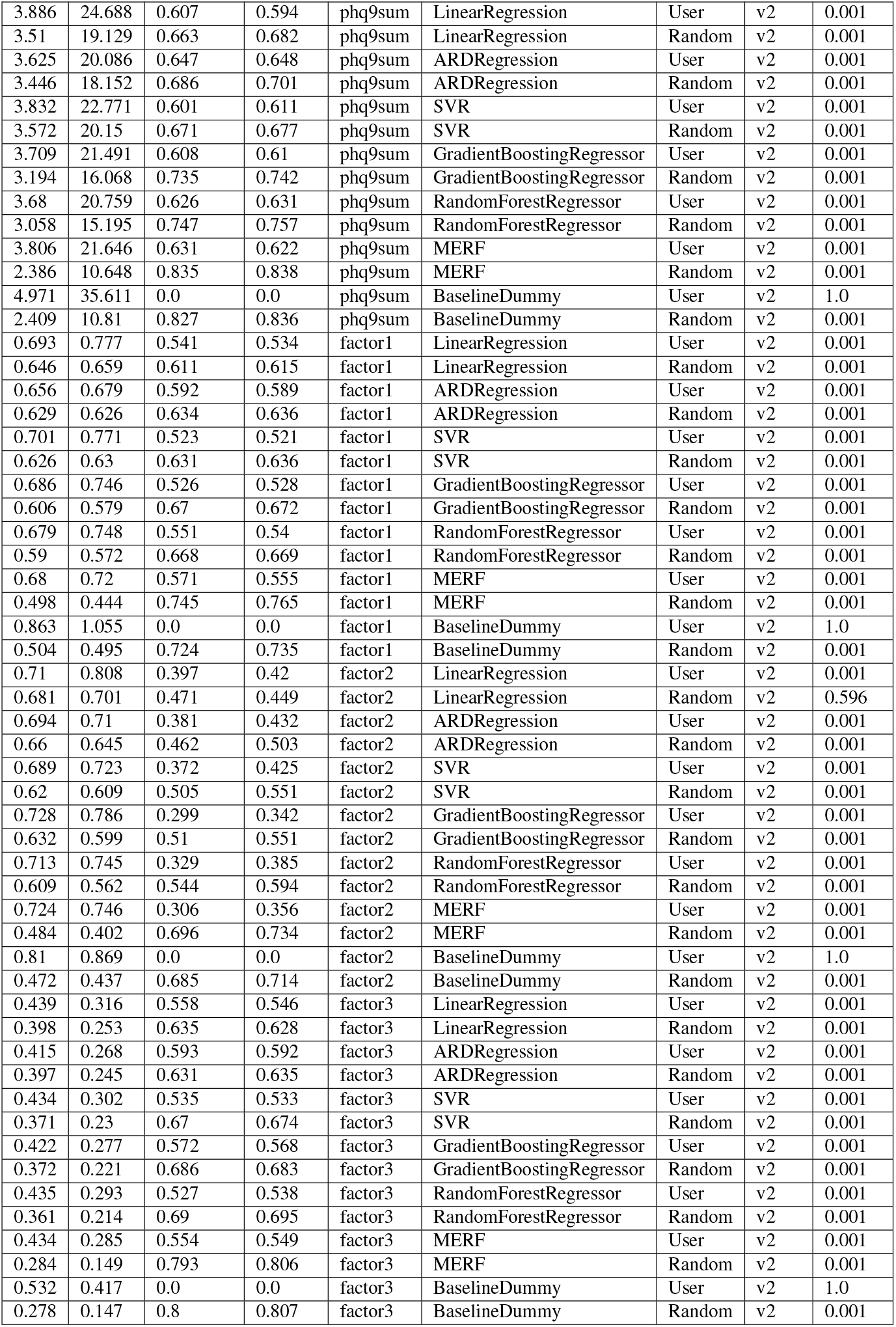

| MAE | MSE | Spearman | Pearson | Target | Estimator | Split | Study | p-value |
| --- | --- | --- | --- | --- | --- | --- | --- | --- |
| 3.178 | 15.953 | 0.674 | 0.683 | phq9sum | MERF | User | v1 | 0.001 |
| 2.216 | 8.749 | 0.833 | 0.84 | phq9sum | MERF | Random | v1 | 0.001 |
| 3.07 | 15.64 | 0.681 | 0.685 | phq9sum | LinearRegression | User | v1 | 0.001 |
| 2.995 | 14.677 | 0.699 | 0.71 | phq9sum | LinearRegression | Random | v1 | 0.001 |
| 3.041 | 15.056 | 0.689 | 0.698 | phq9sum | ARDRegression | User | v1 | 0.001 |
| 3.0 | 14.689 | 0.7 | 0.709 | phq9sum | ARDRegression | Random | v1 | 0.001 |
| 3.172 | 16.547 | 0.661 | 0.666 | phq9sum | SVR | User | v1 | 0.001 |
| 2.922 | 14.45 | 0.715 | 0.719 | phq9sum | SVR | Random | v1 | 0.001 |
| 3.102 | 15.665 | 0.673 | 0.683 | phq9sum | GradientBoostingRegressor | User | v1 | 0.001 |
| 2.871 | 13.626 | 0.722 | 0.735 | phq9sum | GradientBoostingRegressor | Random | v1 | 0.001 |
| 3.17 | 16.464 | 0.658 | 0.666 | phq9sum | RandomForestRegressor | User | v1 | 0.001 |
| 2.761 | 12.961 | 0.738 | 0.749 | phq9sum | RandomForestRegressor | Random | v1 | 0.001 |
| 4.341 | 29.606 | 0.0 | 0.0 | phq9sum | BaselineDummy | User | v1 | 1.0 |
| 2.508 | 11.897 | 0.763 | 0.774 | phq9sum | BaselineDummy | Random | v1 | 0.001 |
| 0.554 | 0.489 | 0.634 | 0.656 | factor1 | MERF | User | v1 | 0.001 |
| 0.455 | 0.363 | 0.737 | 0.764 | factor1 | MERF | Random | v1 | 0.001 |
| 0.561 | 0.505 | 0.642 | 0.647 | factor1 | LinearRegression | User | v1 | 0.001 |
| 0.549 | 0.47 | 0.657 | 0.675 | factor1 | LinearRegression | Random | v1 | 0.001 |
| 0.555 | 0.479 | 0.648 | 0.666 | factor1 | ARDRegression | User | v1 | 0.001 |
| 0.549 | 0.468 | 0.658 | 0.674 | factor1 | ARDRegression | Random | v1 | 0.001 |
| 0.578 | 0.549 | 0.586 | 0.607 | factor1 | SVR | User | v1 | 0.001 |
| 0.514 | 0.452 | 0.671 | 0.691 | factor1 | SVR | Random | v1 | 0.001 |
| 0.548 | 0.49 | 0.635 | 0.657 | factor1 | GradientBoostingRegressor | User | v1 | 0.001 |
| 0.525 | 0.449 | 0.671 | 0.693 | factor1 | GradientBoostingRegressor | Random | v1 | 0.001 |
| 0.56 | 0.504 | 0.622 | 0.646 | factor1 | RandomForestRegressor | User | v1 | 0.001 |
| 0.513 | 0.445 | 0.673 | 0.695 | factor1 | RandomForestRegressor | Random | v1 | 0.001 |
| 0.702 | 0.866 | 0.0 | 0.0 | factor1 | BaselineDummy | User | v1 | 1.0 |
| 0.505 | 0.492 | 0.652 | 0.665 | factor1 | BaselineDummy | Random | v1 | 0.001 |
| 0.68 | 0.685 | 0.407 | 0.496 | factor2 | MERF | User | v1 | 0.001 |
| 0.492 | 0.41 | 0.677 | 0.744 | factor2 | MERF | Random | v1 | 0.001 |
| 0.656 | 0.662 | 0.43 | 0.521 | factor2 | LinearRegression | User | v1 | 0.001 |
| 0.641 | 0.631 | 0.46 | 0.552 | factor2 | LinearRegression | Random | v1 | 0.001 |
| 0.651 | 0.65 | 0.438 | 0.532 | factor2 | ARDRegression | User | v1 | 0.001 |
| 0.64 | 0.63 | 0.457 | 0.553 | factor2 | ARDRegression | Random | v1 | 0.001 |
| 0.699 | 0.754 | 0.358 | 0.443 | factor2 | SVR | User | v1 | 0.001 |
| 0.601 | 0.58 | 0.516 | 0.599 | factor2 | SVR | Random | v1 | 0.001 |
| 0.675 | 0.687 | 0.402 | 0.498 | factor2 | GradientBoostingRegressor | User | v1 | 0.001 |
| 0.633 | 0.605 | 0.484 | 0.577 | factor2 | GradientBoostingRegressor | Random | v1 | 0.001 |
| 0.688 | 0.716 | 0.384 | 0.474 | factor2 | RandomForestRegressor | User | v1 | 0.001 |
| 0.6 | 0.567 | 0.528 | 0.614 | factor2 | RandomForestRegressor | Random | v1 | 0.001 |
| 0.803 | 0.909 | 0.0 | 0.0 | factor2 | BaselineDummy | User | v1 | 1.0 |
| 0.513 | 0.479 | 0.652 | 0.694 | factor2 | BaselineDummy | Random | v1 | 0.001 |
| 0.361 | 0.213 | 0.607 | 0.6 | factor3 | MERF | User | v1 | 0.001 |
| 0.249 | 0.116 | 0.802 | 0.806 | factor3 | MERF | Random | v1 | 0.001 |
| 0.345 | 0.205 | 0.622 | 0.617 | factor3 | LinearRegression | User | v1 | 0.001 |
| 0.337 | 0.194 | 0.646 | 0.641 | factor3 | LinearRegression | Random | v1 | 0.001 |
| 0.343 | 0.2 | 0.632 | 0.627 | factor3 | ARDRegression | User | v1 | 0.001 |
| 0.338 | 0.195 | 0.639 | 0.64 | factor3 | ARDRegression | Random | v1 | 0.001 |
| 0.37 | 0.234 | 0.56 | 0.555 | factor3 | SVR | User | v1 | 0.001 |
| 0.306 | 0.171 | 0.696 | 0.696 | factor3 | SVR | Random | v1 | 0.001 |
| 0.349 | 0.208 | 0.617 | 0.611 | factor3 | GradientBoostingRegressor | User | v1 | 0.001 |
| 0.325 | 0.182 | 0.672 | 0.671 | factor3 | GradientBoostingRegressor | Random | v1 | 0.001 |
| 0.358 | 0.219 | 0.598 | 0.59 | factor3 | RandomForestRegressor | User | v1 | 0.001 |
| 0.315 | 0.177 | 0.688 | 0.682 | factor3 | RandomForestRegressor | Random | v1 | 0.001 |
| 0.454 | 0.331 | 0.0 | 0.0 | factor3 | BaselineDummy | User | v1 | 1.0 |
| 0.271 | 0.142 | 0.753 | 0.758 | factor3 | BaselineDummy | Random | v1 | 0.001 |
| 3.886 | 24.688 | 0.607 | 0.594 | phq9sum | LinearRegression | User | v2 | 0.001 |
| 3.51 | 19.129 | 0.663 | 0.682 | phq9sum | LinearRegression | Random | v2 | 0.001 |
| 3.625 | 20.086 | 0.647 | 0.648 | phq9sum | ARDRegression | User | v2 | 0.001 |
| 3.446 | 18.152 | 0.686 | 0.701 | phq9sum | ARDRegression | Random | v2 | 0.001 |
| 3.832 | 22.771 | 0.601 | 0.611 | phq9sum | SVR | User | v2 | 0.001 |
| 3.572 | 20.15 | 0.671 | 0.677 | phq9sum | SVR | Random | v2 | 0.001 |
| 3.709 | 21.491 | 0.608 | 0.61 | phq9sum | GradientBoostingRegressor | User | v2 | 0.001 |
| 3.194 | 16.068 | 0.735 | 0.742 | phq9sum | GradientBoostingRegressor | Random | v2 | 0.001 |
| 3.68 | 20.759 | 0.626 | 0.631 | phq9sum | RandomForestRegressor | User | v2 | 0.001 |
| 3.058 | 15.195 | 0.747 | 0.757 | phq9sum | RandomForestRegressor | Random | v2 | 0.001 |
| 3.806 | 21.646 | 0.631 | 0.622 | phq9sum | MERF | User | v2 | 0.001 |
| 2.386 | 10.648 | 0.835 | 0.838 | phq9sum | MERF | Random | v2 | 0.001 |
| 4.971 | 35.611 | 0.0 | 0.0 | phq9sum | BaselineDummy | User | v2 | 1.0 |
| 2.409 | 10.81 | 0.827 | 0.836 | phq9sum | BaselineDummy | Random | v2 | 0.001 |
| 0.693 | 0.777 | 0.541 | 0.534 | factor1 | LinearRegression | User | v2 | 0.001 |
| 0.646 | 0.659 | 0.611 | 0.615 | factor1 | LinearRegression | Random | v2 | 0.001 |
| 0.656 | 0.679 | 0.592 | 0.589 | factor1 | ARDRegression | User | v2 | 0.001 |
| 0.629 | 0.626 | 0.634 | 0.636 | factor1 | ARDRegression | Random | v2 | 0.001 |
| 0.701 | 0.771 | 0.523 | 0.521 | factor1 | SVR | User | v2 | 0.001 |
| 0.626 | 0.63 | 0.631 | 0.636 | factor1 | SVR | Random | v2 | 0.001 |
| 0.686 | 0.746 | 0.526 | 0.528 | factor1 | GradientBoostingRegressor | User | v2 | 0.001 |
| 0.606 | 0.579 | 0.67 | 0.672 | factor1 | GradientBoostingRegressor | Random | v2 | 0.001 |
| 0.679 | 0.748 | 0.551 | 0.54 | factor1 | RandomForestRegressor | User | v2 | 0.001 |
| 0.59 | 0.572 | 0.668 | 0.669 | factor1 | RandomForestRegressor | Random | v2 | 0.001 |
| 0.68 | 0.72 | 0.571 | 0.555 | factor1 | MERF | User | v2 | 0.001 |
| 0.498 | 0.444 | 0.745 | 0.765 | factor1 | MERF | Random | v2 | 0.001 |
| 0.863 | 1.055 | 0.0 | 0.0 | factor1 | BaselineDummy | User | v2 | 1.0 |
| 0.504 | 0.495 | 0.724 | 0.735 | factor1 | BaselineDummy | Random | v2 | 0.001 |
| 0.71 | 0.808 | 0.397 | 0.42 | factor2 | LinearRegression | User | v2 | 0.001 |
| 0.681 | 0.701 | 0.471 | 0.449 | factor2 | LinearRegression | Random | v2 | 0.596 |
| 0.694 | 0.71 | 0.381 | 0.432 | factor2 | ARDRegression | User | v2 | 0.001 |
| 0.66 | 0.645 | 0.462 | 0.503 | factor2 | ARDRegression | Random | v2 | 0.001 |
| 0.689 | 0.723 | 0.372 | 0.425 | factor2 | SVR | User | v2 | 0.001 |
| 0.62 | 0.609 | 0.505 | 0.551 | factor2 | SVR | Random | v2 | 0.001 |
| 0.728 | 0.786 | 0.299 | 0.342 | factor2 | GradientBoostingRegressor | User | v2 | 0.001 |
| 0.632 | 0.599 | 0.51 | 0.551 | factor2 | GradientBoostingRegressor | Random | v2 | 0.001 |
| 0.713 | 0.745 | 0.329 | 0.385 | factor2 | RandomForestRegressor | User | v2 | 0.001 |
| 0.609 | 0.562 | 0.544 | 0.594 | factor2 | RandomForestRegressor | Random | v2 | 0.001 |
| 0.724 | 0.746 | 0.306 | 0.356 | factor2 | MERF | User | v2 | 0.001 |
| 0.484 | 0.402 | 0.696 | 0.734 | factor2 | MERF | Random | v2 | 0.001 |
| 0.81 | 0.869 | 0.0 | 0.0 | factor2 | BaselineDummy | User | v2 | 1.0 |
| 0.472 | 0.437 | 0.685 | 0.714 | factor2 | BaselineDummy | Random | v2 | 0.001 |
| 0.439 | 0.316 | 0.558 | 0.546 | factor3 | LinearRegression | User | v2 | 0.001 |
| 0.398 | 0.253 | 0.635 | 0.628 | factor3 | LinearRegression | Random | v2 | 0.001 |
| 0.415 | 0.268 | 0.593 | 0.592 | factor3 | ARDRegression | User | v2 | 0.001 |
| 0.397 | 0.245 | 0.631 | 0.635 | factor3 | ARDRegression | Random | v2 | 0.001 |
| 0.434 | 0.302 | 0.535 | 0.533 | factor3 | SVR | User | v2 | 0.001 |
| 0.371 | 0.23 | 0.67 | 0.674 | factor3 | SVR | Random | v2 | 0.001 |
| 0.422 | 0.277 | 0.572 | 0.568 | factor3 | GradientBoostingRegressor | User | v2 | 0.001 |
| 0.372 | 0.221 | 0.686 | 0.683 | factor3 | GradientBoostingRegressor | Random | v2 | 0.001 |
| 0.435 | 0.293 | 0.527 | 0.538 | factor3 | RandomForestRegressor | User | v2 | 0.001 |
| 0.361 | 0.214 | 0.69 | 0.695 | factor3 | RandomForestRegressor | Random | v2 | 0.001 |
| 0.434 | 0.285 | 0.554 | 0.549 | factor3 | MERF | User | v2 | 0.001 |
| 0.284 | 0.149 | 0.793 | 0.806 | factor3 | MERF | Random | v2 | 0.001 |
| 0.532 | 0.417 | 0.0 | 0.0 | factor3 | BaselineDummy | User | v2 | 1.0 |
| 0.278 | 0.147 | 0.8 | 0.807 | factor3 | BaselineDummy | Random | v2 | 0.001 |

| Study | Sub-Set | Data Points | Subjects | Features |
| --- | --- | --- | --- | --- |
| V1 | EMA | 3007 | 541 | 16 |
| V1 | Communication | 3007 | 541 | 27 |
| V1 | Mobility | 3007 | 541 | 14 |
| V2 | EMA | 1159 | 276 | 16 |
| V2 | Activity | 1159 | 276 | 37 |
| V2 | Communication | 1159 | 276 | 36 |
| V2 | Weather | 1159 | 276 | 39 |

## Bibliography

1. Deborah S. Hasin, Aaron L. Sarvet, Jacquelyn L. Meyers, Tulshi D. Saha, W. June Ruan, Malka Stohl, and Bridget F. Grant. Epidemiology of adult DSM-5 major depressive disorder and its specifiers in the United States. JAMA Psychiatry, 75(4), 2018. ISSN 2168622X. doi: 10.1001/jamapsychiatry.2017.4602.

2. Jean Pierre Lépine and Mike Briley. The increasing burden of depression. Neuropsychiatric Disease and Treatment, 7(SUPPL.), 2011. ISSN 11782021. doi: 10.2147/NDT.S19617.

3. Harold W. Neighbors, Steven J. Trierweiler, Briggett C. Ford, and Jordana R. Muroff. Racial differences in DSM diagnosis using a semi-structured instrument: The importance of clinical judgment in the diagnosis of African Americans. Journal of Health and Social Behavior, 44 (3), 2003. ISSN 00221465. doi: 10.2307/1519777.

4. Astha Singhal, Yu Yu Tien, and Renee Y. Hsia. Racial-ethnic disparities in opioid prescriptions at emergency department visits for conditions commonly associated with prescription drug abuse. PLoS ONE, 11(8), 2016. ISSN 19326203. doi: 10.1371/journal.pone.0159224.

5. John Torous, Mathew V. Kiang, Jeanette Lorme, and Jukka Pekka Onnela. New tools for new research in psychiatry: A scalable and customizable platform to empower data driven smartphone research. JMIR Mental Health, 3(2), 4 2016. ISSN 23687959. doi: 10.2196/mental.5165.

6. Jukka Pekka Onnela and Scott L. Rauch. Harnessing Smartphone-Based Digital Phenotyping to Enhance Behavioral and Mental Health, 6 2016. ISSN 1740634X.

7. Valeria De Angel, Serena Lewis, Katie White, Carolin Oetzmann, Daniel Leightley, Emanuela Oprea, Grace Lavelle, Faith Matcham, Alice Pace, David C. Mohr, Richard Dobson, and Matthew Hotopf. Digital health tools for the passive monitoring of depression: a systematic review of methods. npj Digital Medicine, 5(1), 12 2022. ISSN 23986352. doi: 10.1038/s41746-021-00548-8.

8. Sohrab Saeb, Mi Zhang, Mary Kwasny, Christopher J. Karr, Konrad Kording, and David C. Mohr. The relationship between clinical, momentary, and sensor-based assessment of depression. In Proceedings of the 2015 9th International Conference on Pervasive Computing Technologies for Healthcare, PervasiveHealth 2015, 2015. doi: 10.4108/icst.pervasivehealth.2015.259034.

9. Afsaneh Doryab, Jun Ki Min, Jason Wiese, John Zimmerman, and Jason Hong. Detection of behavior change in people with depression. In AAAI Workshop - Technical Report, volume WS-14-08, 2014.

10. Cláudia T. Moraes, Trinitat Cambras, Antoni Diez-Noguera, Regina Schimitt, Giovana Dantas, Rosa Levandovski, and Maria P. Hidalgo. A new chronobiological approach to discriminate between acute and chronic depression using peripheral temperature, rest-activity, and light exposure parameters. BMC Psychiatry, 13, 2013. ISSN 1471244X. doi: 10.1186/1471-244X-13-77.

11. Anastasiya Slyepchenko, Olivia R. Allega, Xiamin Leng, Luciano Minuzzi, Maha M. Eltayebani, Matthew Skelly, Roberto B. Sassi, Claudio N. Soares, Sidney H. Kennedy, and Benicio N. Frey. Association of functioning and quality of life with objective and subjective measures of sleep and biological rhythms in major depressive and bipolar disorder. Australian and New Zealand Journal of Psychiatry, 53(7), 2019. ISSN 14401614. doi: 10.1177/0004867419829228.

12. Jian Cao, Anh Lan Truong, Sophia Banu, Asim A. Shah, Ashutosh Sabharwal, and Nidal Moukaddam. Tracking and predicting depressive symptoms of adolescents using smartphone-based self-reports, parental evaluations, and passive phone sensor data: Development and usability study. JMIR Mental Health, 7(1), 2020. ISSN 23687959. doi: 10.2196/14045.

13. Ilya Baryshnikov, Talayeh Aledavood, Tom Rosenström, Roope Heikkilä, Richard Darst, Kirsi Riihimäki, Outi Saleva, Jesper Ekelund, and Erkki Isometsä. Relationship between daily rated depression symptom severity and the retrospective self-report on PHQ-9: A prospective ecological momentary assessment study on 80 psychiatric outpatients. Journal of Affective Disorders, 324, 2023. ISSN 15732517. doi: 10.1016/j.jad.2022.12.127.

14. Fabian Wahle, Tobias Kowatsch, Elgar Fleisch, Michael Rufer, and Steffi Weidt. Mobile sensing and support for people with depression: A pilot trial in the wild. JMIR mHealth and uHealth, 4(3), 2016. ISSN 22915222. doi: 10.2196/mhealth.5960.

15. Akane Sano, Sara Taylor, Andrew W. McHill, Andrew J.K. Phillips, Laura K. Barger, Elizabeth Klerman, and Rosalind Picard. Identifying objective physiological markers and modifiable behaviors for self-reported stress and mental health status using wearable sensors and mobile phones: Observational study. Journal of Medical Internet Research, 20(6), 2018. ISSN 14388871. doi: 10.2196/jmir.9410.

16. Rui Wang, Weichen Wang, Alex daSilva, Jeremy F. Huckins, William M. Kelley, Todd F. Heatherton, and Andrew T. Campbell. Tracking Depression Dynamics in College Students Using Mobile Phone and Wearable Sensing. Proceedings of the ACM on Interactive, Mobile, Wearable and Ubiquitous Technologies, 2(1), 2018. doi: 10.1145/3191775.

17. Kennedy Opoku Asare, Isaac Moshe, Yannik Terhorst, Julio Vega, Simo Hosio, Harald Baumeister, Laura Pulkki-Råback, and Denzil Ferreira. Mood ratings and digital biomarkers from smartphone and wearable data differentiates and predicts depression status: A longitudinal data analysis. Pervasive and Mobile Computing, 83, 2022. ISSN 15741192. doi: 10.1016/j.pmcj.2022.101621.

18. Shweta Ware, Chaoqun Yue, Reynaldo Morillo, Jin Lu, Chao Shang, Jinbo Bi, Jayesh Kamath, Alexander Russell, Athanasios Bamis, and Bing Wang. Predicting Depressive Symptoms Using Smartphone Data. Smart Health, 2019. doi: 10.1016/j.smhl.xxxx.xx.xxx.

19. Kennedy Opoku Asare, Yannik Terhorst, Julio Vega, Ella Peltonen, Eemil Lagerspetz, and Denzil Ferreira. Predicting depression from smartphone behavioral markers using machine learning methods, hyperparameter optimization, and feature importance analysis: Exploratory study. JMIR mHealth and uHealth, 9(7), 7 2021. ISSN 22915222. doi: 10.2196/26540.

20. Paola Pedrelli, Szymon Fedor, Asma Ghandeharioun, Esther Howe, Dawn F. Ionescu, Dar-ian Bhathena, Lauren B. Fisher, Cristina Cusin, Maren Nyer, Albert Yeung, Lisa Sangermano, David Mischoulon, Johnathan E. Alpert, and Rosalind W. Picard. Monitoring Changes in Depression Severity Using Wearable and Mobile Sensors. Frontiers in Psychiatry, 11, 12 2020. ISSN 16640640. doi: 10.3389/fpsyt.2020.584711.

21. Robert A. Lewis, Asma Ghandeharioun, Szymon Fedor, Paola Pedrelli, Rosalind Picard, and David Mischoulon. Mixed Effects Random Forests for Personalised Predictions of Clinical Depression Severity. arXiv, 1 2023.

22. Vincent Holstein, Habiballah Rahimi-Eichi, Daniel Emden, Lara Gutfleisch, Alexander Refisch, Janik Goltermann, Ramona Leenings, Nils Winter, Tilo Kircher, Igor Nenadí, Ronny Redlich, Elisabeth Johanna Leehr, Katharina Dohm, Justin Baker, Udo Dannlowski, Nils Opel, and Tim Hahn. Remote monitoring of depression severity: A machine learning approach. medRxiv, 2023. doi: 10.1101/2023.08.22.23294431.

23. Md Sabbir Ahmed and Nova Ahmed. A Fast and Minimal System to Identify Depression Using Smartphones: Explainable Machine Learning–Based Approach. JMIR Formative Research, 7, 2023. ISSN 2561326X. doi: 10.2196/28848.

24. Abhishek Pratap, Ava Homiar, Luke Waninger, Calvin Herd, Christine Suver, Joshua Volponi, Joaquin A. Anguera, and Pat Areán. Real-world behavioral dataset from two fully remote smartphone-based randomized clinical trials for depression. Scientific Data, 9(1), 2022. ISSN 20524463. doi: 10.1038/s41597-022-01633-7.

25. Martin Papenberg and Gunnar W. Klau. Using anticlustering to partition data sets into equivalent parts. Psychological Methods, 26(2), 2021. ISSN 1082989X. doi: 10.1037/met0000301.

26. William Revelle. Package ‘psych’ - Procedures for Psychological, Psychometric, and Personality Research. Technical report, Northwestern University, 2023.

27. Yves Rosseel. Lavaan: An R package for structural equation modeling. Journal of Statistical Software, 48, 2012. ISSN 15487660. doi: 10.18637/jss.v048.i02.

28. Robert C. MacCallum, Michael W. Browne, and Hazuki M. Sugawara. Power analysis and determination of sample size for covariance structure modeling. Psychological Methods, 1 (2), 1996. ISSN 1082989X. doi: 10.1037/1082-989X.1.2.130.

29. Li Tze Hu and Peter M. Bentler. Cutoff criteria for fit indexes in covariance structure analysis: Conventional criteria versus new alternatives. Structural Equation Modeling, 6(1), 1999. ISSN 10705511. doi: 10.1080/10705519909540118.

30. Ramona Leenings, Nils Ralf Winter, Lucas Plagwitz, Vincent Holstein, Jan Ernsting, Kelvin Sarink, Lukas Fisch, Jakob Steenweg, Leon Kleine-Vennekate, Julian Gebker, Daniel Emden, Dominik Grotegerd, Nils Opel, Benjamin Risse, Xiaoyi Jiang, Udo Dannlowski, and Tim Hahn. PHOTONAI-A Python API for rapid machine learning model development. PLoS ONE, 16(July), 7 2021. ISSN 19326203. doi: 10.1371/journal.pone.0254062.

31. Fabian Pedregosa, Gael Varoquaux, Alexandre Gramfort, Vincent Michel, Bertrand Thirion, Olivier Grisel, Mathieu Blondel, Peter Prettenhofer, Ron Weiss, Vincent Dubourg, Jake Vanderplas, Alexandre Passos, David Cournapeau, Matthieu Brucher, Matthieu Perrot, and Édouard Duchesnay. Scikit-learn: Machine learning in Python. Journal of Machine Learning Research, 12, 2011. ISSN 15324435.

32. Ahlem Hajjem, François Bellavance, and Denis Larocque. Mixed-effects random forest for clustered data. Journal of Statistical Computation and Simulation, 84(6), 2014. ISSN 00949655. doi: 10.1080/00949655.2012.741599.

33. Scott M Lundberg and Su-In Lee. A Unified Approach to Interpreting Model Predictions. In Advances in Neural Information Processing Systems, 2017.

34. Andrew Gelman, John B Carlin, Hal S Stern, David B Dunson, Aki Vehtari, Donald B Rubin, John Carlin, Hal Stern, Donald Rubin, and David Dunson. Bayesian Data Analysis - Third Edition. Chapman and Hall/CRC, third edition edition, 2014.

35. Daniel Lewandowski, Dorota Kurowicka, and Harry Joe. Generating random correlation matrices based on vines and extended onion method. Journal of Multivariate Analysis, 100 (9), 2009. ISSN 0047259X. doi: 10.1016/j.jmva.2009.04.008.

36. John K. Kruschke. Doing Bayesian data analysis: A tutorial with R, JAGS, and Stan, second edition. Academic Press, second edition edition, 2014. doi: 10.1016/B978-0-12-405888-0.09999-2.

37. Radford M Neal. Probabilistic inference using Markov chain Monte Carlo methods. Technical report, University of Toronto, Toronto, 1993.

38. Paul Christian Bürkner. brms: An R package for Bayesian multilevel models using Stan. Journal of Statistical Software, 80, 2017. ISSN 15487660. doi: 10.18637/jss.v080.i01.

39. Víctor Manuel López-Guerra, Carla López-Núñez, Silvia L. Vaca-Gallegos, and Pablo V. Torres-Carrión. Psychometric Properties and Factor Structure of the Patient Health Questionnaire-9 as a Screening Tool for Depression Among Ecuadorian College Students. Frontiers in Psychology, 13, 2022. ISSN 16641078. doi: 10.3389/fpsyg.2022.813894.

40. Yang Wang, Lijuan Liang, Zhenyuan Sun, Rongxun Liu, Yange Wei, Shisan Qi, Qiao Ke, and Fei Wang. Factor structure of the patient health questionnaire-9 and measurement invariance across gender and age among Chinese university students. Medicine (United States), 102(1), 2023. ISSN 15365964. doi: 10.1097/MD.0000000000032590.

41. Linh Gia Vu, Linh Khanh Le, Anh Vu Trong Dam, Son Hoang Nguyen, Thuc Thi Minh Vu, Trang Thu Hong Trinh, Anh Linh Do, Ngoc Minh Do, Trang Huyen Le, Carl Latkin, Roger C.M. Ho, and Cyrus S.H. Ho. Factor Structures of Patient Health Questionnaire-9 Instruments in Exploring Depressive Symptoms of Suburban Population. Frontiers in Psychiatry, 13, 2022. ISSN 16640640. doi: 10.3389/fpsyt.2022.838747.

42. Diogo Lamela, Cátia Soreira, Paula Matos, and Ana Morais. Systematic review of the factor structure and measurement invariance of the patient health questionnaire-9 (PHQ-9) and validation of the Portuguese version in community settings. Journal of Affective Disorders, 276, 2020. ISSN 15732517. doi: 10.1016/j.jad.2020.06.066.

43. Dror Ben-Zeev, Emily A. Scherer, Rui Wang, and Haiyi Xie. Next-Generation psychiatric assessment: using smartphone sensors to monitor behavior and mental health. Psychiatric Rehabilitation Journal, 38(3), 2015. ISSN 1095158X. doi: 10.1037/prj0000130.

44. Marcus J.H. Huibers, L. Esther de Graaf, Frenk P.M.L. Peeters, and Arnoud Arntz. Does the weather make us sad? Meteorological determinants of mood and depression in the general population. Psychiatry Research, 180(2-3), 2010. ISSN 01651781. doi: 10.1016/j.psychres.2009.09.016.

45. Johannes C. Eichstaedt, Robert J. Smith, Raina M. Merchant, Lyle H. Ungar, Patrick Crutchley, Daniel Preoţiuc-Pietro, David A. Asch, and H. Andrew Schwartz. Facebook language predicts depression in medical records. Proceedings of the National Academy of Sciences of the United States of America, 115(44), 2018. ISSN 10916490. doi: 10.1073/pnas.1802331115.

46. Caitlin A. Stamatis, Jonah Meyerhoff, Tingting Liu, Garrick Sherman, Harry Wang, Tony Liu, Brenda Curtis, Lyle H. Ungar, and David C. Mohr. Prospective associations of textmessage-based sentiment with symptoms of depression, generalized anxiety, and social anxiety. Depression and Anxiety, 39(12), 2022. ISSN 15206394. doi: 10.1002/da.23286.

47. Eric L. Ross, Kelly L. Zuromski, Ben Y. Reis, Matthew K. Nock, Ronald C. Kessler, and Jordan W. Smoller. Accuracy Requirements for Cost-effective Suicide Risk Prediction among Primary Care Patients in the US. JAMA Psychiatry, 78(6):642–650, 6 2021. ISSN 2168622X. doi: 10.1001/jamapsychiatry.2021.0089.

